# Complement drives PNH red cell hemolysis independently of inflammasome activation

**DOI:** 10.64898/2026.07.20.26358486

**Authors:** Nikhil Ranjan, Michael A. Cole, Gloria F Gerber, Daniel Flores-Guerrero, Shruti Chaturvedi, Robert A. Brodsky

## Abstract

Paroxysmal Nocturnal Hemoglobinuria (PNH) is characterized by hemolysis due to the loss of GPI-anchored complement regulators. While terminal complement inhibitors improve survival, the precise intracellular mechanisms driving the destruction of PNH erythrocytes remain controversial. A recently proposed model suggests PNH cells undergo an inflammatory programmed cell death (“spectosis”) driven by an NLRP3-Caspase-8 signaling cascade. Here, we use a whole packed cell lysis approach to map the cytoskeletal degradation of primary erythrocytes across a 22-patient PNH cohort. Our data show that membrane attack complex (MAC) pore formation drives targeted β-spectrin fragmentation, which correlates with rapid intracellular potassium (K^+^) efflux. Notably, when probing these primary patient samples, we detected a complete absence of the NLRP3 protein and found no functional evidence of Caspase-8 activation during MAC pore formation. Furthermore, caspase inhibition did not alter cytoskeletal degradation or K^+^ efflux. Instead, our data demonstrate that MAC-induced membrane perforation permits a rapid influx of calcium, which activates calpain, the dominant calcium-dependent protease in erythrocytes. Rather than an inflammatory cascade, this calcium-dependent calpain activity executes the degradation of β-spectrin. These findings challenge current models of PNH hemolysis. We show that the destruction of PNH erythrocytes is a consequence of the MAC-calcium-calpain axis, rather than an inflammatory programmed cell death event. Consequently, therapeutic strategies aimed at targeting the inflammasome or caspase signaling will likely offer no clinical benefit for PNH patients.

**Graphical abstract:** 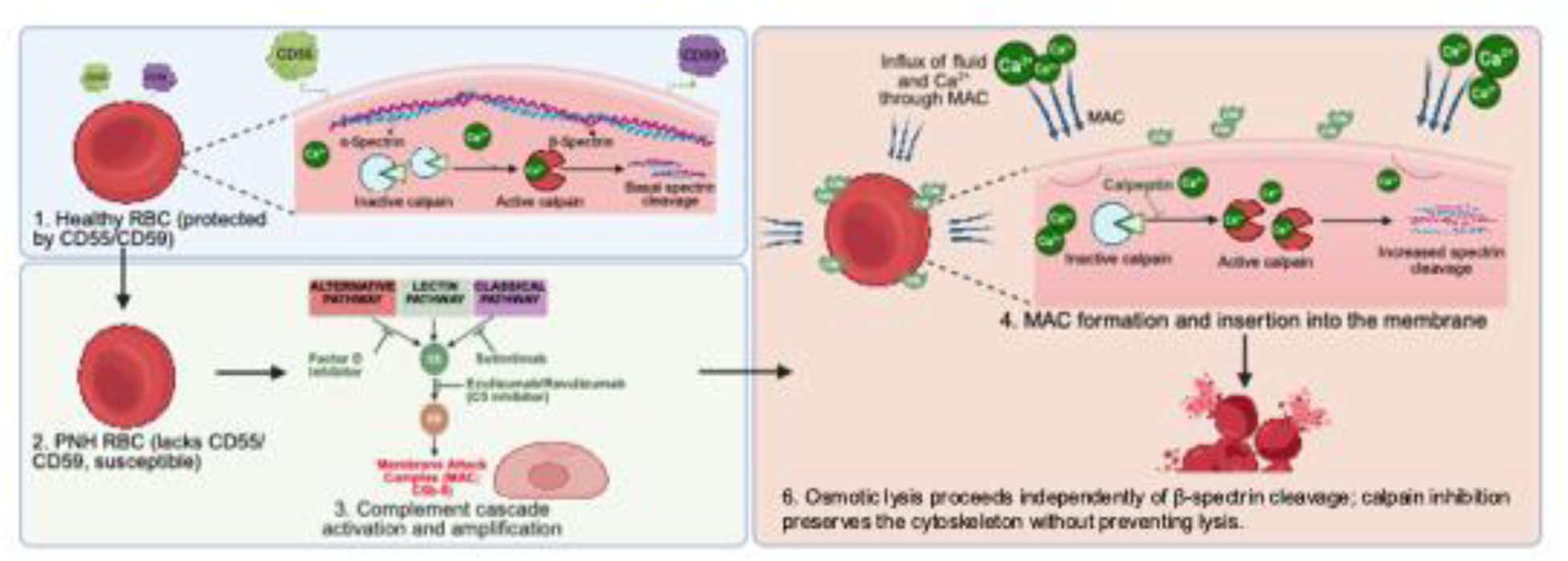

## Introduction

Paroxysmal Nocturnal Hemoglobinuria (PNH) is an acquired hematopoietic stem cell disorder caused by somatic mutations in the PIGA gene, which results in the loss of glycosylphosphatidylinositol (GPI)-anchored proteins [1–6]. The disease is defined by intravascular hemolysis and thrombosis, primarily due to the absence of the complement regulators CD55 and CD59. Lacking these membrane regulators, PNH erythrocytes are highly susceptible to membrane attack complex (MAC)-mediated lysis [7, 8]. Although terminal complement inhibitors like eculizumab and ravulizumab have drastically improved patient survival, they do not cure the disease [1, 4, 9–11]. Patients frequently experience “breakthrough” intravascular hemolysis during physiological stress, often driven by C5 bypass mechanisms [1, 10, 12]. Furthermore, because PNH erythrocytes also lack the C3 convertase regulator CD55, cells surviving C5 blockade become heavily opsonized with C3 fragments, driving extravascular hemolysis [1, 9, 13, 14]. Even with clinically effective complement blockade, PNH erythrocytes still exhibit a reduced lifespan. This ongoing fragility indicates that MAC-induced damage extends beyond simple membrane perforation, likely involving the underlying structural architecture of the red cell [15].

A healthy erythrocyte maintains its shape and flexibility through the submembrane cytoskeleton, a dense lattice of α- and β-spectrin anchored to the lipid bilayer [16–19]. While structural defects in this network are well-documented in hereditary disorders like spherocytosis [16], we know surprisingly little about what happens to the RBC cytoskeleton during complement-mediated destruction in PNH.

A recent study proposed that PNH erythrocytes undergo “spectosis”, a form of programmed cell death driven by inflammatory signaling [20]. In this model, complement activation is hypothesized to trigger an intracellular “mini-NLRP3” inflammasome, which recruits and activates Caspase-8 to directly cleave β-spectrin. This implies that inhibiting NLRP3 or Caspase-8 could mitigate hemolysis. However, this pathway has not been validated in primary cells from a broad clinical cohort. Understanding whether PNH cells die through inflammatory signaling or direct biophysical failure has important therapeutic implications.

Here, we investigated the structural failure of PNH red cells using a whole packed cell lysis technique designed to retain the complete erythroid proteome, including soluble fragments often lost during ghost preparation. Our results challenge the “spectosis” model. We show that while PNH erythrocytes undergo targeted β-spectrin fragmentation linked to rapid potassium (K^+^) efflux, this degradation is entirely independent of NLRP3 or Caspase-8. Across a clinical cohort of 22 PNH patients, we found an absolute absence of the NLRP3 machinery and no functional evidence of caspase activity in erythrocytes. Instead, MAC pore formation drives an influx of calcium that activates calpain, the dominant resident erythrocyte protease [21–24]. It is this calcium-dependent calpain activity, not an inflammatory cascade, that executes β-spectrin degradation. These findings suggest that the mechanism of PNH red cell destruction differs fundamentally from the spectosis model and that targeting programmed cell death pathways may not benefit PNH patients.

## Materials and methods

### Patient Samples and Erythrocyte Preparation

Whole blood was collected following informed consent from 22 patients with a confirmed PNH clone of >1% across multiple lineages, as determined by clinical flow cytometry. Erythrocytes were isolated by centrifugation at 500 x g for 5 minutes at 25°C. Plasma and the buffy coat were discarded, and the remaining red cells were washed three times in sterile PBS. After the final wash, erythrocytes were resuspended to a 2% hematocrit in GVB^0^ (containing no calcium or magnesium Mg²⁺), which was then supplemented with 3 mM MgEGTA to yield a final working concentration of 3 mM free magnesium. This buffer selectively blocks the classical and lectin pathways while permitting alternative pathway (AP) amplification. Patients who had received red blood cell transfusions within 4 months prior to sample collection were included in flow cytometric analyses (with appropriate gating for CD59-negative populations) but were strictly excluded from all bulk in vitro hemolysis and immunoblotting assays to prevent confounding from wild-type donor erythrocytes. This study was approved by the Institutional Review Board at Johns Hopkins University and was conducted in accordance with the Declaration of Helsinki. All patients provided written informed consent.

### Flow Cytometry and Clone Size Determination

To quantify the GPI-deficient erythrocytes (PNH erythrocyte clone size) and stratify patient samples, flow cytometric analysis was performed on peripheral blood samples as described previously [25]. Briefly, washed erythrocytes were resuspended in FACS buffer (HBSS supplemented with 1% BSA) and stained with an APC conjugated CD59 antibody (Clone MEM-43, Thermo Scientific). Data acquisition was performed using a Cytoflex S flow cytometer, and analysis was conducted using FlowJo v10 software. Within the gated erythrocyte population, cells were assessed for CD59 surface expression to identify the Type III PNH clone. These experimentally derived clone sizes, utilized to stratify the experimental cohorts for functional and structural analyses, were cross-referenced and independently corroborated with the patients’ official clinical diagnostic flow cytometry records to ensure absolute cohort accuracy prior to all functional assays. For hemolysis assays, patients were stratified as large (>50%) or small (<50%) clones. For immunoblotting analyses of spectrin cleavage, we focused on patients with very large (>90%) and very small (<5%) clones to maximize the contrast between affected and unaffected populations.

### Pharmacological Inhibition

Erythrocytes were pre-incubated with various inhibitors at 30°C for 30 minutes prior to addition of 20% serum. Conditions included terminal complement blockade with eculizumab (338 nM/50µg/mL; clinical grade), AP-specific blockade with a Factor D inhibitor (FDi, 1 μM), and classical pathway inhibition with sutimlimab (41 nM/ 6µg/mL; clinical grade). To evaluate programmed cell death pathways, we utilized the pan-caspase inhibitor Z-VAD-FMK (10 μM, MedChemExpress), the caspase-8 specific inhibitor Z-IETD-FMK (10 μM, MedChemExpress), and the NLRP3 inhibitor MCC950 (10 μM, Enzo Lifesciences). Cells were additionally pre-incubated with the calcium-binding domain calpain inhibitor PD150606 (50µM, Sigma) or the proteasome inhibitor epoxomicin (1 µM, Sigma) for 30min at 37°C prior to complement or ionophore challenge. Serum samples were pre-treated with complement inhibitors at 4°C for 30 minutes. Vehicle controls were treated with equivalent volumes of buffer (DMSO <0.1%). To evaluate the role of calcium-dependent proteases, cells were treated with the targeted calpain inhibitor calpeptin (25–100 µM, Sigma). To bypass the complement cascade and directly induce global calcium overload, erythrocytes were stimulated with the calcium ionophore A23187 (8 µM, Sigma).

### RBC lysis assay and quantification

Alternative pathway-mediated lysis was initiated by adding ABO-matched normal human serum (NHS) to the 2% erythrocyte suspension to a final concentration of 20%. To selectively induce and isolate alternative pathway amplification, the assay was performed in acidified GVB^0^-3mM MgEGTA buffer (pH 6.4). Following a 2-hour incubation at 37°C, samples were centrifuged at 500 x g for 5 minutes at 4°C. The cell-free supernatant was transferred to a 96-well U-bottom plate, and hemoglobin release was quantified by measuring absorbance at 541 nm (BMG CLARIOstar, Germany). Spontaneous lysis (background) was determined using erythrocytes in buffer alone, and 100% lysis was established using distilled water. The percentage of hemolysis for each condition was calculated using the following formula: Hemolysis (%) = (Absorbance Sample−Absorbance Background) × 100/(Absorbance 100% Lysis−Absorbance Background)

### Cell culture

Wild-type TF-1 cells were obtained from the American Type Culture Collection (ATCC) and maintained in RPMI-1640 medium supplemented with 10% fetal bovine serum (FBS) and 2 ng/mL GM-CSF at 37°C in a 5% CO₂ atmosphere. To generate a GPI-deficient model, TF-1 cells were treated with 1 nM aerolysin (AL) for 7 to 10 days to select for PIGA-null populations. The complete loss of GPI-anchored surface proteins was confirmed via flow cytometry. In functional validation assays, aerolysin also served as a positive control for GPI-dependent, MAC-independent pore formation.

### Intracellular Potassium (K^+^) Efflux Kinetics

Intracellular K^+^ levels were monitored using the cell-permeable fluorescent indicator IPG-4 AM (Ion Biosciences). TF-1*^WT^*and TF-1*^PIGAnull^* cells were washed in HBSS +10 mM HEPES and loaded with 3.5 μM IPG-4 AM for 90 minutes at 37°C in the dark. Cells were washed twice and resuspended in K^+^ free assay buffer (10 mM HEPES, 145 mM NaCl, 0.15 mM CaCl_2_, and 0.5 mM MgCl_2_). For kinetic measurements, 10^5^ cells/well were seeded in a 96-well black-bottom microplate. Baseline fluorescence (Ex: 525 nm/ Em: 545 nm) was recorded for 2 minutes at 37°C. Cells were then stimulated with 10% NHS or heat-inactivated serum (HIS), and fluorescence was recorded every minute for 60 minutes. Valinomycin (20 µM) was used as a positive control to establish maximal K^+^ depletion. Kinetic values were normalized to the initial baseline (F/F0); a decrease in fluorescence indicates K^+^ efflux through MAC-induced membrane pores.

### Erythrocyte Ghost Preparation, Whole Cell Lysis, and Immunoblotting

To assess complement-mediated structural damage, erythrocytes were incubated with 20% normal human serum in GVB++ buffer (supplemented with 2 mM Ca²⁺ and 2 mM Mg²⁺) to permit the necessary extracellular calcium influx upon MAC pore formation. Following the incubation, to capture the complete erythroid proteome and retain soluble breakdown products, erythrocytes were pelleted at 20,000 x g for 10 minutes at 4°C. The supernatant was aspirated, and pellets were lysed directly in 1X NuPAGE LDS Sample Buffer (Invitrogen) supplemented with 50 mM DTT, 1X Halt Protease Inhibitor Cocktail (Thermo Scientific), and 50 mM Tris-HCl. Samples were vigorously vortexed and heated at 70°C for 10 minutes. For comparative subcellular fractionation, erythrocyte ghosts were prepared by lysing cells in hypotonic buffer (5 mM sodium phosphate, pH 8.0) containing protease inhibitors. Ghosts were pelleted at 20,000 x g for 20 minutes and washed iteratively until the pellet was entirely white, after which the membrane fractions were solubilized in LDS sample buffer.

To maintain a constant representation of the initial cellular input, lysates were loaded volumetrically (representing ∼1.3 µL of initial packed RBCs per lane) and resolved on NuPAGE 4–12% Bis-Tris gels (Invitrogen) using MOPS running buffer. Proteins were transferred to low-fluorescence PVDF membranes (Immobilon FL, Merck). To account for loading variations without relying on internal ratio methods, membranes were stained with REVERT 700 Total Protein Stain (LI-COR Biosciences) for 5 minutes and imaged to provide a definitive Total Protein Stain (TPS) normalization factor for densitometric quantification.

Membranes were blocked in Intercept TBS Blocking Buffer (LI-COR) for 1 hour. For structural analysis, membranes were probed overnight at 4°C with primary antibodies against Spectrin β I (B-1, 1:1000; Santa Cruz) or Spectrin α I (B-12, 1:1000; Santa Cruz). To assess inflammasome presence, membranes were probed with NLRP3 (Clone Cryo-2, 1:2000; Adipogen) targeting the PYD domain, and NLRP3 (1:1000; CST) targeting the NACHT domain. THP-1 cell lysates served as a positive control for NLRP3 expression. Following washes, membranes were incubated with near-infrared secondary antibodies (1:20,000; LI-COR) and visualized using the Odyssey DLx Imaging System (LI-COR). All densitometric measurements were internally normalized to the total protein signal (Total Protein Stain) in each lane to ensure accurate quantification despite variations in protein recovery.

### Statistical Analysis

Data are presented as the mean ± standard deviation (SD) from n=3 independent experiments unless otherwise mentioned. Statistical significance was determined using one-way analysis of variance (ANOVA) followed by Dunnett’s multiple comparisons test to evaluate differences between experimental conditions and controls. A p-value of <0.05 was considered statistically significant. All statistical analyses were performed using GraphPad Prism software (version 10).

## Results

### Complement-mediated hemolysis of PNH erythrocytes is independent of caspase or NLRP3 inflammasome activation

To evaluate complement-mediated hemolysis, we optimized buffer conditions for AP activation. Standard saline did not support lysis, but magnesium only buffer and calcium chelation (GVB^0^-3mM MgEGTA) drove maximal erythrocyte destruction (Supplemental Figure S1). We used this buffer for subsequent complement assays.

We next subjected primary PNH erythrocytes to this assay using 20% ABO-matched normal human serum. Active serum induced hemolysis that correlated directly with the patient’s GPI-deficient cell burden. Patients with smaller clones (<50%, n=11) showed proportionally less hemolysis (Figure 1B), while those with a large Type III clone (>50%, n=11) exhibited lysis approaching their total clone size (Figure 1C). Heat-inactivated (HI) serum prevented this lysis entirely. Treatment with the terminal C5 inhibitor eculizumab or the AP-specific Factor D inhibitor (FDi) rescued the cells in both cohorts.

**Figure 1.**
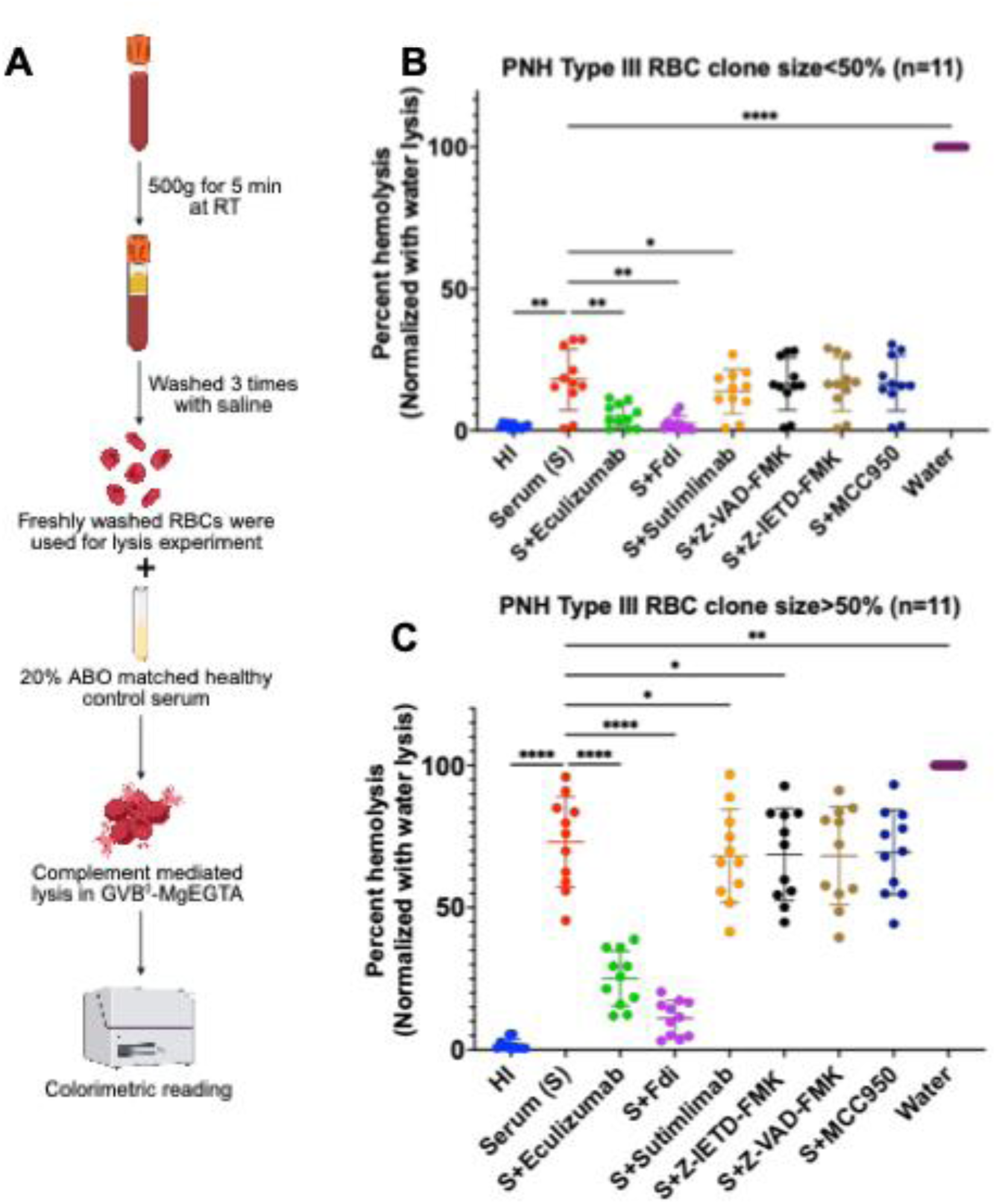
Complement-mediated hemolysis of PNH erythrocytes is independent of caspase and NLRP3 inflammasome signaling. **(A)** Schematic representation of the in vitro alternative pathway (AP)-mediated hemolysis assay. Primary erythrocytes from PNH patients were isolated, washed, and challenged with 20% ABO-matched normal human serum in an acidified GVB^0^-3mM MgEGTA buffer optimized for AP amplification. Hemolysis was quantified via colorimetric measurement of free hemoglobin release. **(B, C)** Percent hemolysis of primary erythrocytes from PNH patients stratified by **(B)** a smaller Type III RBC clone size (<50%, n=11) and **(C)** a large Type III clone size (>50%, n=11). Erythrocytes/serum were pre-incubated with the indicated pharmacological inhibitors for 30 minutes prior to complement challenge. Terminal complement blockade (Eculizumab, 338 nM) and AP-specific blockade (Factor D inhibitor [Fdi], 1 µM) completely rescued cells from active serum-induced lysis. Conversely, classical pathway inhibition (Sutimlimab, 41 nM), Caspase-8 inhibition (Z-IETD-FMK, 10 µM), pan-caspase inhibition (Z-VAD-FMK, 10 µM), and NLRP3 blockade (MCC950, 10 µM) failed to confer protection. Heat-inactivated (HI) serum served as a negative control. Distilled water served as a positive control for 100% lysis. Hemolysis is normalized to distilled water lysis. Data points represent individual patient samples. Horizontal lines and error bars denote mean ± SD. Statistical significance was determined by repeated measures one-way ANOVA with Dunnett’s multiple comparisons test; *p < 0.05, **p < 0.01, ****p < 0.0001 compared to active serum control.

To confirm the mechanistic specificity of this complement-driven lysis, we utilized the classical pathway inhibitor (sutimlimab) as a negative control. To further test whether this hemolysis requires active programmed signaling, we inhibited caspases and the inflammasome. The pan-caspase inhibitor (Z-VAD-FMK) and the NLRP3 inhibitor (MCC950) completely failed to protect PNH erythrocytes from serum-induced hemolysis. While specific Caspase-8 inhibition (Z-IETD-FMK) and classical pathway blockade (sutimlimab) registered a statistically significant reduction in hemolysis, these effects were biologically negligible (<5% mean difference) and are likely attributable to intrinsic clone size variations within the patient cohorts. These treatments failed to provide the profound, near-total cellular rescue achieved by terminal complement or AP blockade (Figure 1B, 1C).

### MAC pore formation mediates rapid intracellular potassium efflux independent of inflammasome activation

To distinguish between direct MAC pore formation and secondary inflammasome cascades, we tracked the real-time kinetics of intracellular potassium (K⁺) efflux using the fluorescent indicator IPG-4 AM. We normalized kinetic data to baseline fluorescence (F/F0) to account for variations in dye loading. Validation with aerolysin [26], a pore-forming toxin requiring GPI-anchored proteins, showed rapid K⁺ efflux in wild-type (WT) TF-1 cells (Figure 2A, 2C), whereas PIGA-null TF-1 cells remained functionally immune and maintained baseline K⁺ levels (Figure 2B, 2D).

**Figure 2.**
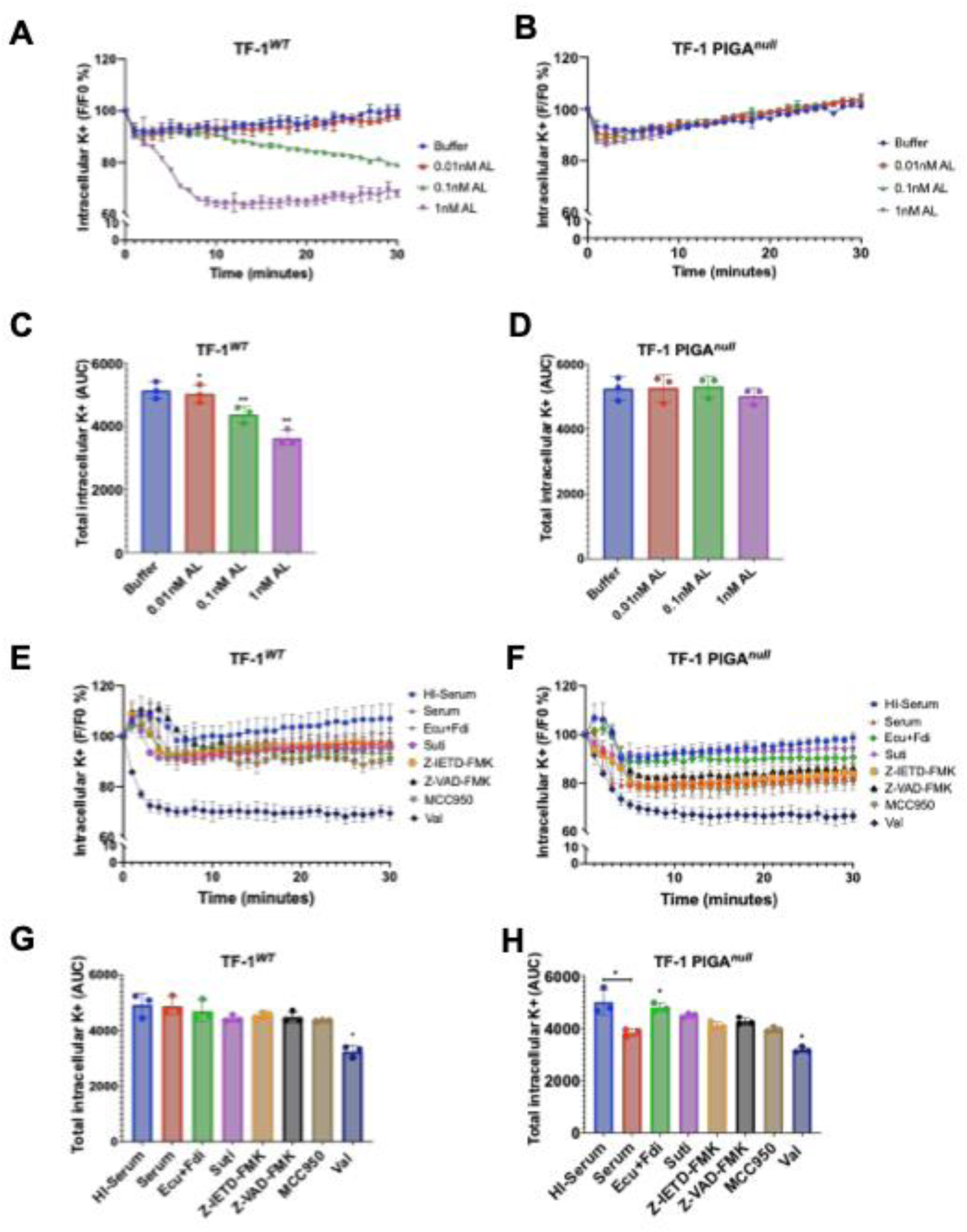
MAC pore formation mediates rapid intracellular potassium (K^+^) efflux independent of secondary inflammasome signaling. (A-B) Assay validation and time-course measurement of intracellular potassium (K⁺) levels in **(A)** TF-1*^WT^* and **(B)** TF-1 PIGA*^null^*cells following treatment with GPI-dependent pore-forming toxin aerolysin (AL) at indicated concentrations (0.01 nM, 0.1 nM, 1 nM) or buffer control. Intracellular K⁺ was monitored using a fluorescent potassium indicator (IPG-4AM) and expressed as the ratio of fluorescence intensity relative to baseline (F/F0 %). Measurements were recorded every minute for 30 minutes. **(C-D)** Quantification of total intracellular K⁺ calculated as area under the curve (AUC) from time-course data in panels A and B for **(C)** TF-1^WT^ and **(D)** TF-1 PIGA*^null^* cells. Data demonstrate dose-dependent K⁺ efflux in TF-1^WT^ cells but not in TF-1PIGA*^null^* cells. **(E–H)** Real-time K^+^ efflux kinetics in response to complement attack. Cells were challenged with 10% active normal human serum (Serum) or heat-inactivated serum (HI-Serum). TF-1 *^WT^* cells resist complement-mediated pore formation due to the retention of cell surface CD59 **(E, G)**. In contrast, TF-1 PIGA*^null^* cells undergo rapid and profound K^+^ depletion upon exposure to active serum **(F, H)**. This K^+^ loss is entirely abrogated by dual complement blockade [Ecu (338 nM) +Fdi (1µM)] and classical pathway inhibition (Suti, 41 nM). Conversely, pharmacological inhibition of Caspase-8 (Z-IETD-FMK, 10 µM), pan-caspases (Z-VAD-FMK, 10 µM), or NLRP3 (MCC950, 10 µM) failed to rescue the cells, providing only a minor blunting of the early efflux phase. Valinomycin (Val, 20µM) served as a positive control for maximal intracellular K^+^ depletion. Kinetic data **(A, B, E, F)** are normalized to initial baseline fluorescence (F/F0 %). Total intracellular K^+^ **(C, D, G, H)** was quantified as the area under the curve (AUC). Data represent the mean ± SD from n=3 independent experiments performed in duplicates. *p < 0.05, **p < 0.01 as determined by One-way ANOVA with Dunnett’s multiple comparisons test, compared to the buffer control (C, D) or active serum (G, H).

When stimulated with 10% active normal human serum, TF-1*^WT^*cells fully resisted complement pore formation due to endogenous CD59 (Figure 2E, 2G). However, TF-1*^PIGA-null^* cells underwent rapid K⁺ efflux upon serum exposure (Figure 2F, 2H). Dual blockade with eculizumab and FDi abrogated the K^+^ loss. Because the K⁺ efflux assay was performed in a buffer containing 0.15 mM Ca²⁺ and 0.5 mM Mg²⁺ to support live-cell fluorescence, classical pathway activation was permitted; hence, inhibition with sutimlimab also provided substantial protection in this assay. Regardless of the initiating pathway of complement-dependent cell killing, we found little evidence that downstream signaling drives this ion dysregulation. While the caspase and NLRP3 inhibitors provided a partial blunting of the initial K⁺ efflux, they did not achieve the near-total rescue seen with complement blockade (Figure 2H). This minor effect in a nucleated cell model suggests that direct MAC insertion, rather than secondary signaling, is the primary driver of rapid K⁺ depletion.

### Mature human erythrocytes lack the NLRP3 inflammasome machinery

The “spectosis” model relies on the assembly of an intracellular NLRP3 inflammasome to drive complement-mediated cell death. To determine if primary, enucleated red blood cells possess this machinery, we assessed NLRP3 protein expression. To ensure comprehensive detection and rule out epitope masking, we utilized two distinct antibodies targeting different structural domains of the protein (the PYD domain and the NACHT domain).

As expected, lysates from THP-1 monocytes provided a robust positive control, displaying clear NLRP3 expression at approximately 120 kDa (Figure 3A, 3B). In contrast, we detected no NLRP3 protein in highly concentrated preparations of either isolated RBC ghosts or whole packed RBCs (Figure 3A, 3B). Importantly, while the Cryo-2 clone exhibited high-molecular-weight off-target binding in the RBC ghost fractions corresponding to the size of spectrin (∼240 kDa), this represents a non-specific technical artifact rather than true protein complexing. Total protein and globin loading controls confirmed the presence of dense erythroid protein in these lanes. To determine whether this reflects a broader absence of inflammasome-associated proteins or is specific to NLRP3, we cross-referenced an independent, deep proteomic resource of the human RBC membrane and cytoplasmic proteome generated from pooled healthy donor blood (>5,200 proteins, ∼88,000 peptide-spectrum matches) [27]. Consistent with our immunoblotting data, NLRP3 was undetectable at either the protein or peptide level in either fraction. Notably, this same dataset identified ASC (PYCARD) and pro-caspase-8 with high confidence (5–7 unique peptides each, FDR q<0.01) at PSM counts comparable to or lower than those supporting the NLRP3-negative call, indicating the assay was sufficiently sensitive to detect proteins of similarly low abundance. These data indicate that mature erythrocytes retain the downstream adaptor and effector components of this pathway (ASC, pro-caspase-8) but specifically lack the upstream nucleating sensor, NLRP3 precluding assembly of a canonical or “mini” NLRP3 inflammasome complex.

**Figure 3.**
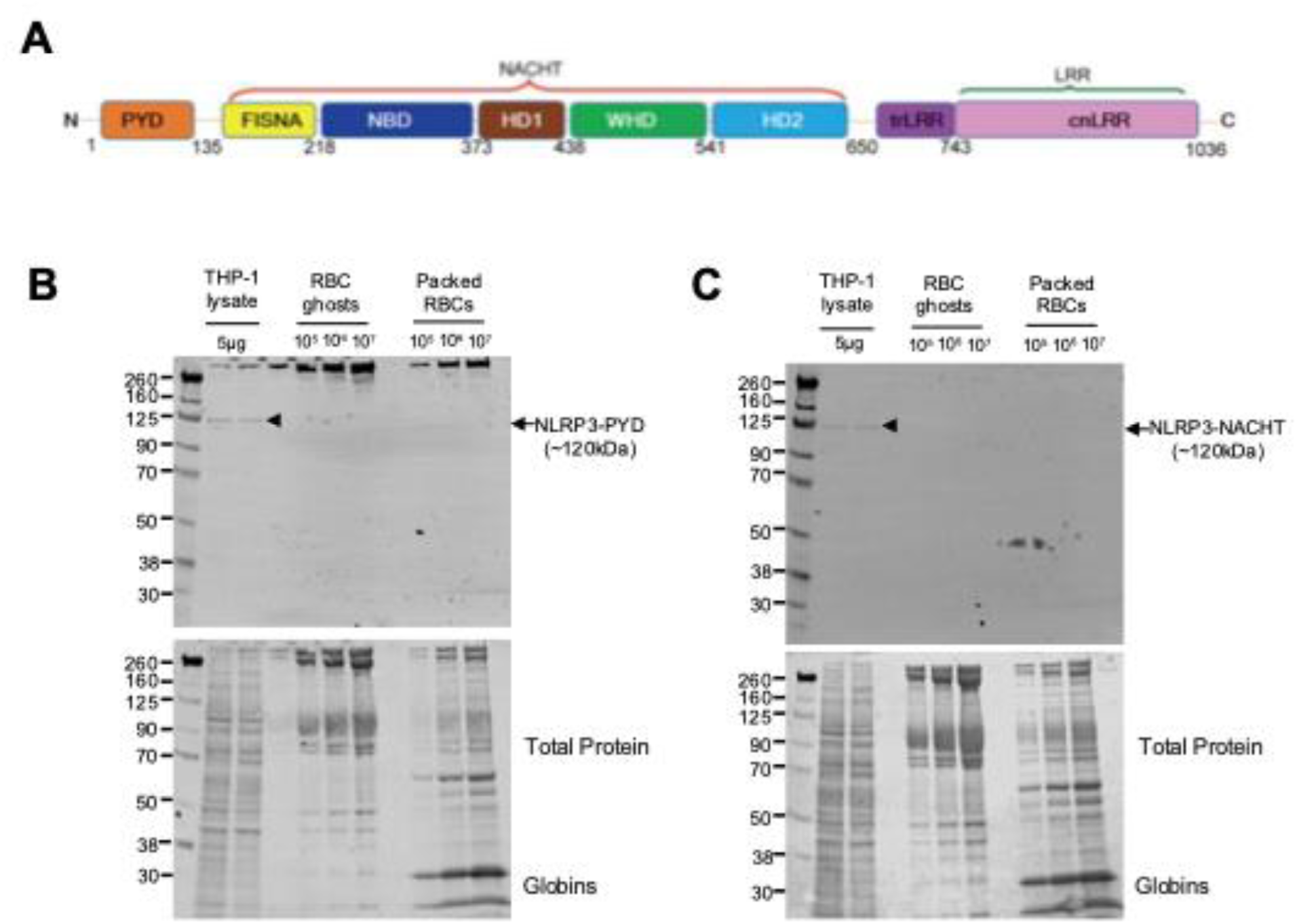
Mature human erythrocytes lack NLRP3 inflammasome structural components. **(A)** Schematic representation of the full-length NLRP3 protein domain structure, highlighting the N-terminal Pyrin domain (PYD), the central NACHT domain, and the C-terminal Leucine-rich repeat (LRR) domain. **(B, C)** Immunoblot analysis of NLRP3 protein expression across nucleated and enucleated cell lysates. To comprehensively evaluate expression and rule out epitope masking, membranes were probed with two highly specific monoclonal antibodies targeting distinct structural regions of the protein: **(B)** the PYD domain (∼120 kDa) and **(C)** the NACHT domain (∼120 kDa). THP-1 monocyte lysates (5 µg) provided a robust positive control for NLRP3 expression (black arrowheads). The full blot was examined across the entire resolvable molecular weight range (∼15–250 kDa), including the ∼70–90 kDa region predicted for a leucine-rich-repeat-truncated ‘mini’ NLRP3 variant retaining the PYD and NACHT domains; no immunoreactive band was detected at this or any other molecular weight in either antibody probe. NLRP3 was absent in highly concentrated preparations of isolated RBC ghosts (10^5^, 10^6^, and 10^7^ cell equivalents) and whole packed RBCs (10^5^, 10^6^, and 10^7^ cell equivalents). The open arrowhead denotes prominent, high-molecular-weight off-target binding (∼260 kDa) observed with the Cryo-2 clone in RBC ghost fractions, consistent with spectrin cross-reactivity. Total protein stain (Revert 700) and prominent globin bands (∼15–30 kDa) serve as loading controls, confirming dense erythroid protein loading in the absence of any detectable NLRP3 signal. Molecular weight markers are indicated in kDa. Blots are representative of n = 3 independent experiments.

### Complement-induced cytoskeletal degradation is targeted to β-spectrin and executed by calpain

We next examined the structural consequences of MAC-induced ion flux on the erythrocyte cytoskeleton. Inhibiting the calcium-dependent protease calpain — using either the active-site inhibitor calpeptin or the calcium-binding domain inhibitor PD150606 — provided no protection against complement- or ionophore-mediated cell death (Figure 4A), indicating that MAC-induced osmotic stress drives lysis independently of calpain activity. However, these same calpain inhibitors uncoupled this primary osmotic death from downstream cytoskeletal degradation, largely preserving the β-spectrin network (Figure 4C, E). This structural preservation by calpeptin was consistent across the concentrations tested (25–100 µM) (Supplementary Figure S4).

**Figure 4.**
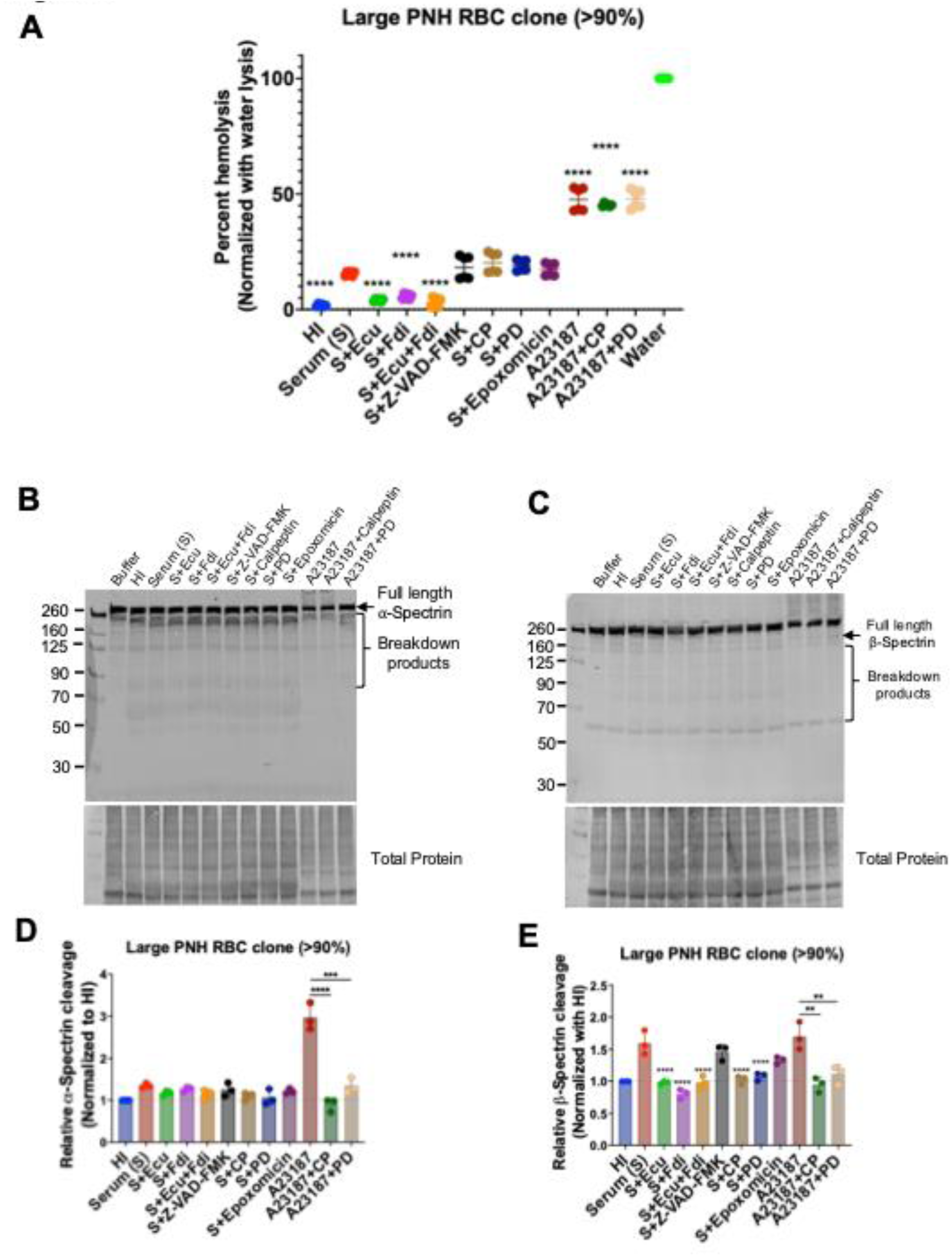
Complement-induced cytoskeletal degradation is uncoupled from hemolysis, highly targeted to β-spectrin, and executed specifically by calpain. (A) Percent hemolysis of primary erythrocytes from patients with large PNH clones (>90%) following challenge with 20% active normal human serum (Serum) or the calcium ionophore A23187 (8 µM). Pharmacological inhibition of the active site of calpain (Calpeptin [CP], 50 µM) or the calcium-binding domain (PD150606 [PD], 50 µM) completely failed to rescue erythrocytes from complement- or ionophore-mediated lysis, demonstrating that osmotic death occurs independently of calpain activity. Generalized proteasome inhibition (Epoxomicin, 1µM) also provided no protection. (B, D) Immunoblot analysis and corresponding densitometric quantification of α-spectrin. Active serum did not induce significant α-spectrin cleavage. Direct calcium overload via A23187 induced α-spectrin cleavage that was abrogated by calpeptin and PD150606. (C, E) Immunoblot analysis and corresponding densitometric quantification of β-spectrin. Active serum induced profound β-spectrin degradation. This targeted cleavage was completely prevented by dual complement blockade (Ecu + Fdi) and by orthogonal calpain inhibitors (CP and PD). Conversely, pan-caspase inhibition (Z-VAD-FMK, 10 µM) and proteasome inhibition (Epoxomicin) failed to protect the cytoskeletal architecture. Lysates were loaded volumetrically (∼1.3 µL packed RBCs per well). Total protein stains (Revert 700) served as loading controls, and all densitometric quantification was internally normalized to the respective total protein signal. Cleavage is expressed relative to the heat-inactivated (HI) serum baseline. Data represent the mean ± SD of n=3 independent experiments. **p < 0.01, ***p < 0.001 ****p < 0.0001 compared to the active serum control by repeated measures one-way ANOVA with Dunnett’s multiple comparisons test.

To capture the complete profile of soluble breakdown products, we analyzed whole erythrocyte lysates rather than isolated membrane ghosts. Purely mechanical cell bursting (water lysis) did not reproduce the β-spectrin cleavage seen with active serum (Figure 4C), indicating that physical membrane rupture alone is not sufficient to drive this cleavage. The proteasome inhibitor epoxomicin similarly failed to prevent the structural collapse (Figure 4C), consistent with a calpain-driven, rather than proteasome-dependent, mechanism.

Active serum induced targeted, partial cleavage of β-spectrin in erythrocytes from patients with large PNH clones (>90%), generating distinct lower-molecular-weight breakdown products (Figure 4C). Importantly, this complement-mediated degradation was undetectable in the small PNH clone cohort (<5%), where the β-spectrin profile remained comparable to baseline controls (Supplemental Figure S3C, S3D).

Full-length α-spectrin, by contrast, remained largely resistant to complement in both the large and small clone cohorts, closely mirroring heat-inactivated controls (Figure 4B, 4D; Supplemental Figure S3A, S3B). This selectivity does not reflect an inherent resistance of α-spectrin to calpain: global calcium overload via the ionophore A23187 induced robust α-spectrin cleavage that was substantially reduced by calpain inhibition (Figure 4B, 4D). Together, this suggests that the localized calcium influx generated by MAC pore formation is sufficient to trigger β-spectrin degradation but falls below the physiological threshold required to cleave α-spectrin.

Together, these data demonstrate that MAC-induced cytoskeletal breakdown in PNH erythrocytes is a calcium/calpain-dependent process that operates independently of inflammasome and caspase cascades.

## Discussion

The hallmark of PNH is the terminal pathway mediated destruction of circulating erythrocytes lacking GPI-anchored complement regulators [1–3, 9, 28]. While terminal C5 inhibitors represent a major clinical milestone, patients remain vulnerable to breakthrough intravascular hemolysis that bypasses the C5 blockade, as well as C3-mediated extravascular hemolysis. Proximal AP inhibitors, such as iptacopan and danicopan, were specifically developed to halt this extravascular destruction. Although these proximal therapies are highly effective, patients can continue to develop persistent or breakthrough anemia driven by ongoing intra- or extravascular hemolytic events [1, 9, 14, 29, 30]. Because clinical complement blockade is rarely absolute, understanding the precise intracellular events that execute the ultimate cytoskeletal collapse following MAC formation remains a critical piece of PNH pathology.

A recent study by Chen et al. proposed that MAC-induced membrane perturbation triggers an NLRP3- and Caspase-8-dependent programmed cell death cascade, termed “spectosis” culminating in β-spectrin cleavage [20]. However, translating these findings to the pathology of PNH requires reconciling a delayed, multi-step programmed signaling model with the rapid, osmotically driven kinetics of true complement-mediated hemolysis. In this study, using an optimized in vitro assay with primary human PNH erythrocytes, we demonstrate that MAC pore formation drives a rapid, physical cellular disruption that operates independently of the inflammasome. Here, we identify the calcium-dependent protease calpain as the primary executioner of cytoskeletal collapse in PNH.

The divergence of our findings from the spectosis model can be attributed to distinct methodological and biophysical constraints. Previous studies reporting delayed, caspase-dependent kinetics heavily utilized standard saline buffers (normal saline), which lack the optimized ion concentrations necessary to sustain the Alternative Pathway amplification loop in vitro (Supplemental Figure S1). By using a magnesium-supplemented, calcium-chelated buffer (3mM-MgEGTA) to robustly drive MAC formation in vitro, our data demonstrate that when terminal complement activation is strongly induced, lysis is rapid, purely physical, and entirely independent of caspase activity. This reflects the universal execution mechanism of PNH, regardless of the initiating complement pathway. Furthermore, previous reports relied on the mouse monoclonal antibody Cryo-2 to detect an erythrocyte-intrinsic “mini-NLRP3” variant [20]. Using rigorous subcellular fractionation designed to optimally preserve the membrane skeleton, we observed significant off-target spectrin binding with the Cryo-2 clone. Additionally, when probed with highly specific, sequence-validated antibodies targeting two distinct domains, we detected complete absence of NLRP3 in both whole-cell and ghost-enriched erythrocyte fractions.

The unique biology of mature erythrocytes constrains, but does not eliminate, their capacity for programmed signaling. Enucleated RBCs lack transcriptional machinery and functional mitochondria, and are therefore incapable of the canonical, Apaf-1-dependent apoptosome amplification loop [31–34]. However, our data, corroborated by an independent proteomic survey of the human RBC proteome [27] indicate that RBCs retain some of the protein machinery downstream of NLRP3, including ASC and pro-caspase-8, consistent with earlier reports that mature erythrocytes express caspase-3 and caspase-8 protein [34]. The deficit we observe is specific to NLRP3 itself, the obligate upstream sensor required to nucleate ASC and recruit caspase-8 in this pathway; without it, ASC and pro-caspase-8 have no scaffold for spectosis-type assembly, regardless of their basal abundance. Because the MAC functions as a non-selective pore, its insertion instead allows a massive influx of extracellular calcium. Foundational biochemical studies have established that calcium influx in erythrocytes preferentially activates resident calpains rather than dormant caspases [33–35].

Our data support a calpain-dependent, rather than caspase-mediated, mechanism of spectrin cleavage during complement-mediated hemolysis. The proteolytic signature observed in our PNH model, specifically the 110 kDa β-spectrin fragment (SBDP110), is a recognized product of calpain cleavage [36, 37]. This conclusion is further supported by our finding that isolated calcium influx via the ionophore A23187 was sufficient to drive widespread spectrin cleavage, which was successfully rescued by the targeted calpain inhibitor calpeptin.

While we observed a minor, partial protective effect from caspase inhibition in our nucleated TF-1 cell model, this signaling axis appears functionally irrelevant in enucleated primary erythrocytes. In our clinical PNH cohort, broad-spectrum caspase inhibition (Z-VAD-FMK) and targeted NLRP3 blockade (MCC950) failed to prevent β-spectrin degradation or erythrocyte hemolysis. Although specific Caspase-8 inhibition (Z-IETD-FMK) and classical pathway blockade (sutimlimab) led to statistically significant reductions in hemolysis in patients with large PNH clones (p < 0.05), the magnitude of this effect (<5% reduction) is biologically negligible compared to the near-complete protection afforded by terminal or proximal (AP) complement blockade (66–85% reduction). Importantly, the failure of pan-caspase and NLRP3 blockade to reach statistical significance further solidifies the conclusion that inflammasome activation is not a major driver of hemolysis in PNH.

Our data reveal a hierarchy of calpain substrate sensitivity during complement attack. While both α- and β-spectrin are fundamentally susceptible to calpain-mediated proteolysis, as demonstrated by the robust cleavage of both chains following global calcium overload with A23187, complement-mediated MAC pore formation selectively degrades β-spectrin while sparing α-spectrin. Whether this selectivity reflects differential calcium thresholds for calpain-mediated cleavage, spatial proximity of β-spectrin to MAC pore insertion sites, or distinct calpain isoform involvement remains to be determined. This observation, supported by Chen et al. [20], highlights an unexplored aspect of calpain substrate selectivity in erythrocytes that needs further investigation.

Identifying calpain as the executioner of MAC-induced collapse raises several important questions. It will be important to investigate the broader degradome of calpain within the erythrocyte during complement attack to determine if other regulatory proteins are simultaneously compromised. This uncoupling between lysis and cytoskeletal injury may also be relevant beyond the acute hemolytic episode. Patients on effective C5 or proximal complement blockade continue to exhibit a shortened erythrocyte lifespan and remain vulnerable to C3-mediated extravascular clearance [1, 9, 13–15], raising the possibility that sublytic, calpain-mediated cytoskeletal injury in surviving cells contributes to their reduced deformability and splenic removal. Whether the degree of β-spectrin cleavage we observed is sufficient to meaningfully affect erythrocyte survival, and whether calpain inhibition could improve deformability in sublytically-attacked cells, remains to be tested. Furthermore, assessing whether this calpain-dominant mechanism is conserved across other complement-mediated hematological disorders will help map shared pathological pathways. This includes not only hemolytic anemias such as Cold Agglutinin Disease (CAD) [38, 39].

In conclusion, our work refines the understanding of complement-mediated erythrocyte death. By demonstrating that PNH red cell destruction relies primarily on the direct physical consequences of the MAC-calcium-calpain axis, we highlight the unique biological constraints of the mature erythrocyte. While inflammasome blockade provides no mechanistic rescue against terminal lysis, future investigations should determine how this distinct, calpain-mediated cytoskeletal fragmentation might alter erythrocyte deformability and influence the extravascular clearance of osmotically stressed cells in vivo.

**Table 1.** Baseline Demographics and Clinical Characteristics of the PNH Cohort.

| Characteristic | Total Cohort (n = 22) |
| --- | --- |
| **Demographics** |  |
| Age — years, median (IQR) [range] | 49.0 (37.0 – 62.0) [20 – 91] |
| Female sex — no. (%) | 13 (59.1%) |
| Race — no. (%) |  |
| White | 18 (81.8%) |
| Black | 2 (9.1%) |
| Asian | 1 (4.5%) |
| White/Hispanic | 1 (4.5%) |
| **Complement-Directed Therapy: no. (%)** |  |
| Ravulizumab monotherapy | 15 (68.2%) |
| Ravulizumab + Danicopan | 3 (13.6%) |
| Iptacopan | 2 (9.1%) |
| Pegcetacoplan | 1 (4.5%) |
| Treatment naïve / None | 1 (4.5%) |
| **PNH Clone Size: median (IQR) [range]** |  |
| Granulocyte clone size — % | 95.3 (86.6 – 99.0) [31.9 – 100.0] |
| RBC clone size — % | 63.8 (35.5 – 97.2) [1.1 – 100.0] |
| Type III RBC clone size — % | 43.2 (21.0 – 89.0) [0.7 – 99.7] |
| **Laboratory Parameters: median (IQR) [range]** |  |
| Hemoglobin — g/dL | 11.9 (9.7 – 13.0) [8.6 – 14.3] |
| Platelet count — $\times 10^3/\mu\text{L}$ | 150 (122 – 204) [32 – 308] |
| Lactate Dehydrogenase (LDH) — U/L | 262 (211 – 341) [161 – 547] |
| Reticulocyte count — cells/$\mu\text{L}$ | 207,100 (118,300 – 266,400) [45,400 – 393,200] |
| Total Bilirubin — mg/dL | 1.1 (0.7 – 2.0) [0.4 – 5.0] |

## Data Availability

All data produced in the present study are available upon reasonable request to the authors.

