## Supplementary figures and images for "Complement drives PNH red cell hemolysis independently of inflammasome activation"

### Supplementary data

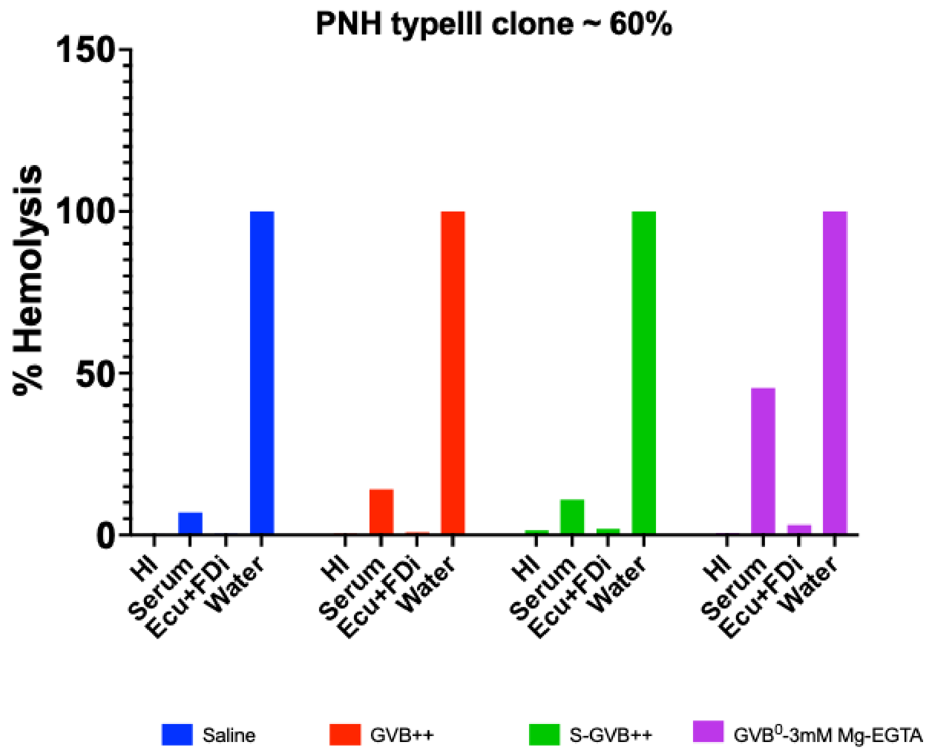

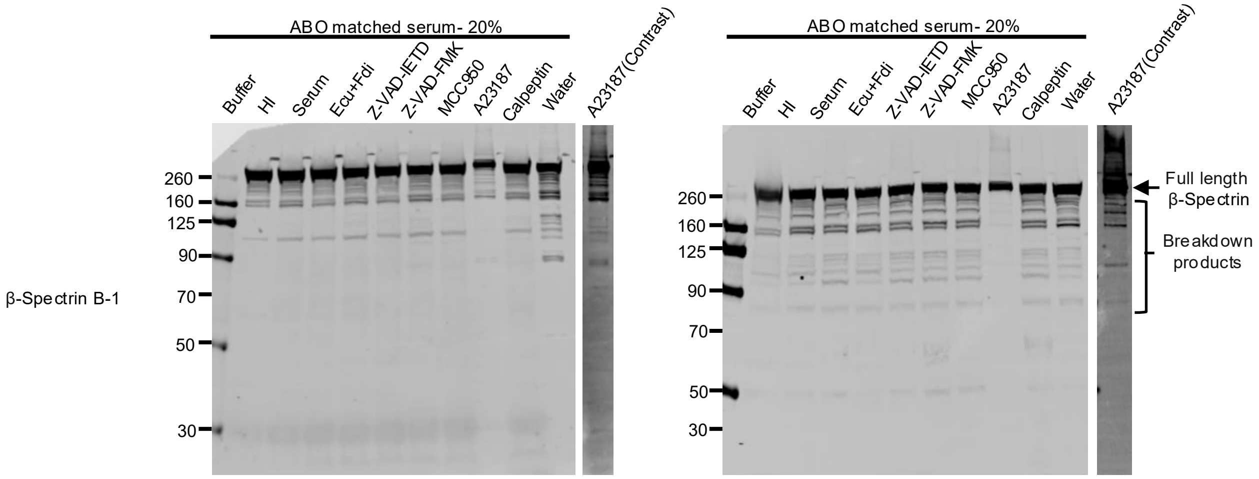

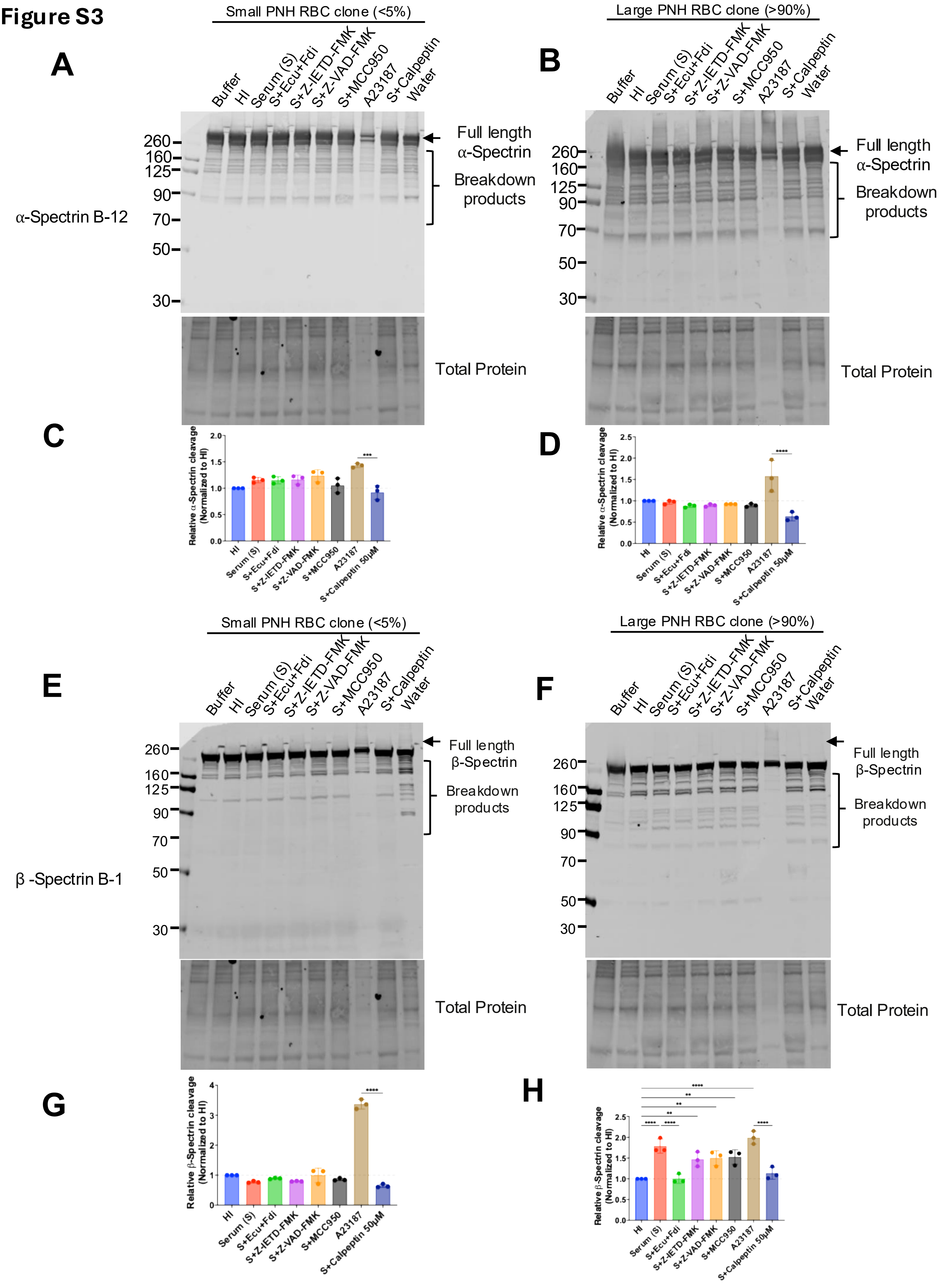

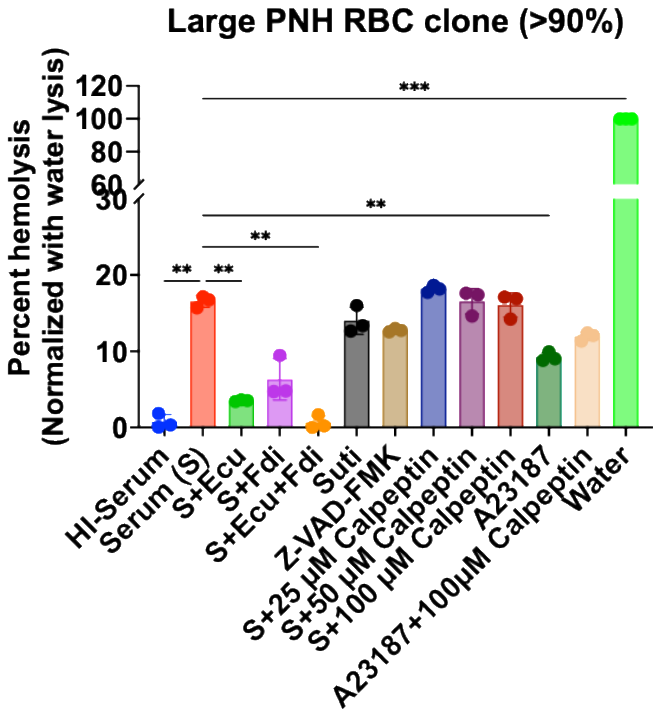
